# Iron status and the risk of sepsis and severe COVID-19: A two-sample Mendelian randomization study

**DOI:** 10.1101/2022.06.02.22275901

**Authors:** Randi Marie Mohus, Helene Flatby, Kristin V. Liyanarachi, Andrew T. DeWan, Erik Solligård, Jan Kristian Damås, Bjørn Olav Åsvold, Lise T. Gustad, Tormod Rogne

## Abstract

**Introduction:** Observational studies have indicated an association between iron status and risk of sepsis and severe COVID-19. However, these findings may be affected by residual confounding, reverse causation.

**Methods:** In a two-sample Mendelian randomization study using inverse variance weighted method, we estimated the effect of genetically-predicted iron biomarkers (serum iron, transferrin saturation (TSAT), total iron binding capacity (TIBC) and ferritin) on risk of sepsis and risk of being hospitalized with COVID-19. For the COVID-19 outcomes we additionally conducted sex-stratified analyses. Weighted median, Weighted mode and MR Egger were used as sensitivity analyses.

**Results:** For risk of sepsis, one standard deviation increase in genetically-predicted serum iron was associated with odds ratio (OR) of 1.14 (95% confidence interval [CI] 1.01 to 1.29, *P*=0.031). The findings were supported in the analyses for transferrin saturation and total iron binding capacity, while the estimate for ferritin was inconclusive. We found a tendency of higher risk of hospitalization with COVID-19 for serum iron; OR 1.29 (CI 0.97–1.72, *P*=0.08), where sex stratified analyses showed OR 1.63 (CI 0.94–2.86, *P*=0.09) for women and OR 1.21 (CI 0.92–1.62, *P*=0.17) for men. Sensitivity analyses supported the main findings and did not suggest bias due to pleiotropy.

**Conclusions:** Our findings suggest a causal effect of genetically-predicted higher iron status and risk of hospitalization due to sepsis and indications of an increased risk of being hospitalized with COVID-19. These findings warrant further studies to assess iron status in relation to severe infections, including the potential of improved management.

## Introduction

Iron is an essential element to various physiological processes, including immune function, metabolism, and erythropoiesis (1, 2). Deviations in iron status (e.g., iron deficiency or iron overload) can have considerable health implications and iron status deviations show substantial sex differences with women more at risk of iron deficiency (1, 3). Iron status can be assessed clinically by using serum iron, transferrin saturation (TSAT), total iron binding capacity (TIBC) and ferritin (3, 4). A growing body of evidence has demonstrated an essential role of systemic and cellular iron-regulating mechanisms in protecting hosts from infections, and most pathogens depend on iron for their pathogenicity (2). Observational studies have indicated an association between iron status and risk of severe infections, where both low iron status (5, 6) and high iron status (7-9) have been linked to increased risk (10, 11). Studies related to COVID-19 found evidence that iron deficiency measured at hospitalization (12), or low serum iron and TSAT but high ferritin (13), were linked to severe COVID-19. On the other hand, excess serum iron, TSAT and lower TIBC (i.e. indication of iron overload) and hyperferritinemia have been associated with critical illness from COVID-19 (14, 15). In a study examining nutritional status in European populations, there were indications of low iron status linked to higher mortality from COVID-19 (16). There is evidence of sex differences in incidence and outcomes of COVID-19 infection (17-19). Few studies have evaluated sex differences in iron status at time of infection. In a small study iron status were lower in female patients when measured at hospitalization due to COVID-19 (15).

A key limitation of observational studies is that they are prone to bias due to confounding and reverse causation. Mendelian randomization (MR) studies can overcome these limitations by using genetic variants associated with the exposures as instrumental variables. Because genetic variants are distributed randomly at conception, the risk of confounding (e.g. from lifestyle factors) and reverse causation (i.e. that the disease affects levels of the exposure) is greatly reduced (20). A recent MR study found a positive association between genetically-predicted high levels of iron biomarkers and risk of sepsis (21), but a more recent set of genetic instruments for iron status has since been published (22). No study has evaluated the role of iron status on the risk of COVID-19 in an MR framework and there is a lack of studies assessing sex differences (23) using sex-stratified MR analyses.

Leveraging data from large genome-wide association studies (GWAS), we aimed to evaluate the association between genetically-predicted iron status biomarkers and risk of being hospitalized with sepsis or COVID-19. In addition, by using sex specific summary-level data on iron status and COVID-19 outcomes, we assessed sex differences in the associations between genetically-predicted iron status and risk of hospitalization due to COVID-19.

## Methods

We performed a two-sample MR study to estimate the effect of genetically-predicted markers of iron status on risk of sepsis and COVID-19 outcomes. None of the iron biomarkers reflect iron status perfectly and iron status in populations is challenging to assess (3, 4). Ferritin is widely used to assess global iron stores but is heavily influenced by inflammation (3, 4). Serum iron is a measure of the fraction of iron that circulates which is readily available and most of it is bound to transferrin. Serum iron is subject to diurnal variation and is affected by fasting status. By measuring the total number of binding sites for iron atoms on transferrin, we calculate the TIBC. TSAT reflects the amount of binding sites on transferrin occupied with iron (calculated as [Serum iron]/[TIBC] %). The normal range is narrow, which is attributed to lower physiological variation than the other iron biomarkers. Low serum iron, low TSAT, low ferritin and high TIBC reflect low iron status. Elevated serum iron, TSAT and ferritin and low TIBC indicate high iron status. The iron in circulation turns over very quickly, especially during infection and inflammation and in clinical conditions with tissue destruction or repeated transfusions (4).

### Genetic instruments for iron status

The exposure of interest was iron status and we ran the analyses for the four iron biomarkers serum iron, TSAT, TIBC and ferritin. The genetic instruments for the iron biomarkers were collected from a GWAS published in 2021 of 246,139 participants of European ancestry (22). The selected single nucleotide polymorphisms (SNPs) used as instruments were strongly associated (p-value <5e-8) with at least one iron biomarker (assumption 1 of MR studies), they should share no common cause with sepsis or COVID-19 (assumption 2), and should only affect the outcome through the risk factor (assumption 3) (20). F statistic above 10 was required for sufficient strength to limit bias due to weak intrumental variables (24). To reduce possible bias due to population stratification, both exposure and outcome cohorts included individuals of European ancestry. Independence between SNPs were ensured by using the LD-reference panel of European populations in 10,000 kb windows and *R*^2^ < 0.01 that is included in the TwoSampleMR (version 0.5.6) package in R (25), and we adjusted for correlation between SNPs using MendelianRandomization (version 0.6.0) in R (version 4.2.1) (26). Sex-specific effects for each biomarker were extracted from the same iron status GWAS using similar precautions for correlation between SNPs (22). We estimated *R*^*2*^ in the TwoSampleMR package and calculated F-statistics using the formula F= ([n-k-1]/k)([ *R*^*2*^*/*1-*R*^*2*^]) (24). The included numbers of SNPs with F-statistics and explained variance of the iron biomarkers is presented for all and separately for men and women, in Supplemental Table S1.

### Genetic susceptibility to sepsis and COVID-19

The genetic susceptibility to sepsis was collected from the IEU OpenGWAS with summary-level data obtained from the UK Biobank which included 10,154 sepsis cases, defined as explicit sepsis (27), and 454,764 controls (25, 28). For COVID-19, we used data from the COVID-19 Host Genetics Initiative (HGI), which is an international collaboration to facilitate COVID-19 genetics research, release 5 (18 Jan 2021). We evaluated two different COVID-19 outcomes: Hospitalized COVID-19 patients (n=4,829) compared with non-hospitalized COVID-19 patients (n=11,816), and hospitalized COVID-19 patients (n=9,986) compared with population-based controls (n=1,877,672) (29). Additionally, we used the sex specific summary-level data on the two COVID-19 outcomes from UKBiobank only, using the NHLBI GRASP catalogue (18 Jun 2021). As with the non-stratified analyses, we used two different COVID-19 outcomes: Hospitalized COVID-19 cases compared with non-hospitalized COVID-19 patients (female cases: n=1,181, controls n=7,586; male cases: 1,703, male controls: n=6,081), and hospitalized with COVID-19 compared to non-hospitalized population (female cases =1,181, controls =248,118; male: cases =1,703, controls =208,248) (30). Unfortunately, we were not able to find sex-stratified summary-level on sepsis.

### MR analyses

The main analysis was the inverse variance weighted (IVW) method which assumes all genetic instruments to be valid (31) and a p-value of 0.05 was used for statistical significance. Three sensitivity analyses were conducted: weighted median, weighted mode, and MR Egger regression. The weighted median orders MR estimates produced by each SNP by their magnitude weighted for their precision and gives an overall MR estimate based on the median value with standard errors estimated by bootstrapping. This method allows for some of the IVs to be invalid (32). The weighted mode assumes that the most common causal effect is consistent with the true causal effect and allows some invalid instruments without biasing the MR estimate (33). MR Egger allows directional pleiotropic effects where some SNPs could be acting on the outcome through another pathway than the exposure of interest, but at the cost of statistical power (34). A consistent effect across these three sensitivity analyses and the IVW analysis suggests that pleiotropy did not bias the IVW estimate.

We used leave-one-out analyses to evaluate whether the IVW estimates were strongly driven by single SNPs (31). Additionally we used PhenoScanner version 2 (35) to check if any of the genetic instruments had important pleiotropic associations. All summary data used in this work are publicly available and with relevant ethical approvals (22, 28, 29), and follow recommendations of reporting MR studies according to STROBE-MR guidelines (36).

## Results

### Sepsis

Genetically-predicted serum iron and TSAT levels were associated with risk of sepsis: Odds ratio (OR) 1.15 (95% confidence interval (CI) 1.01 –1.29, *P*= 0.03) for each standard deviation (SD) (7.76 μmol/L) increase in serum iron; and OR 1.12 (95% CI 1.02 – 1.23, *P*=0.01) per SD increase in TSAT (13.25 %) (Figure 1). The direction of effect for TIBC showed evidence of lower TIBC (i.e. indicating increased iron status) being associated with sepsis OR 0.94 (95% CI 0.87 – 1.01, *P*=0.09). Ferritin showed inconclusive results. The sensitivity analyses supported the findings from the IVW analyses.

**Figure 1:**
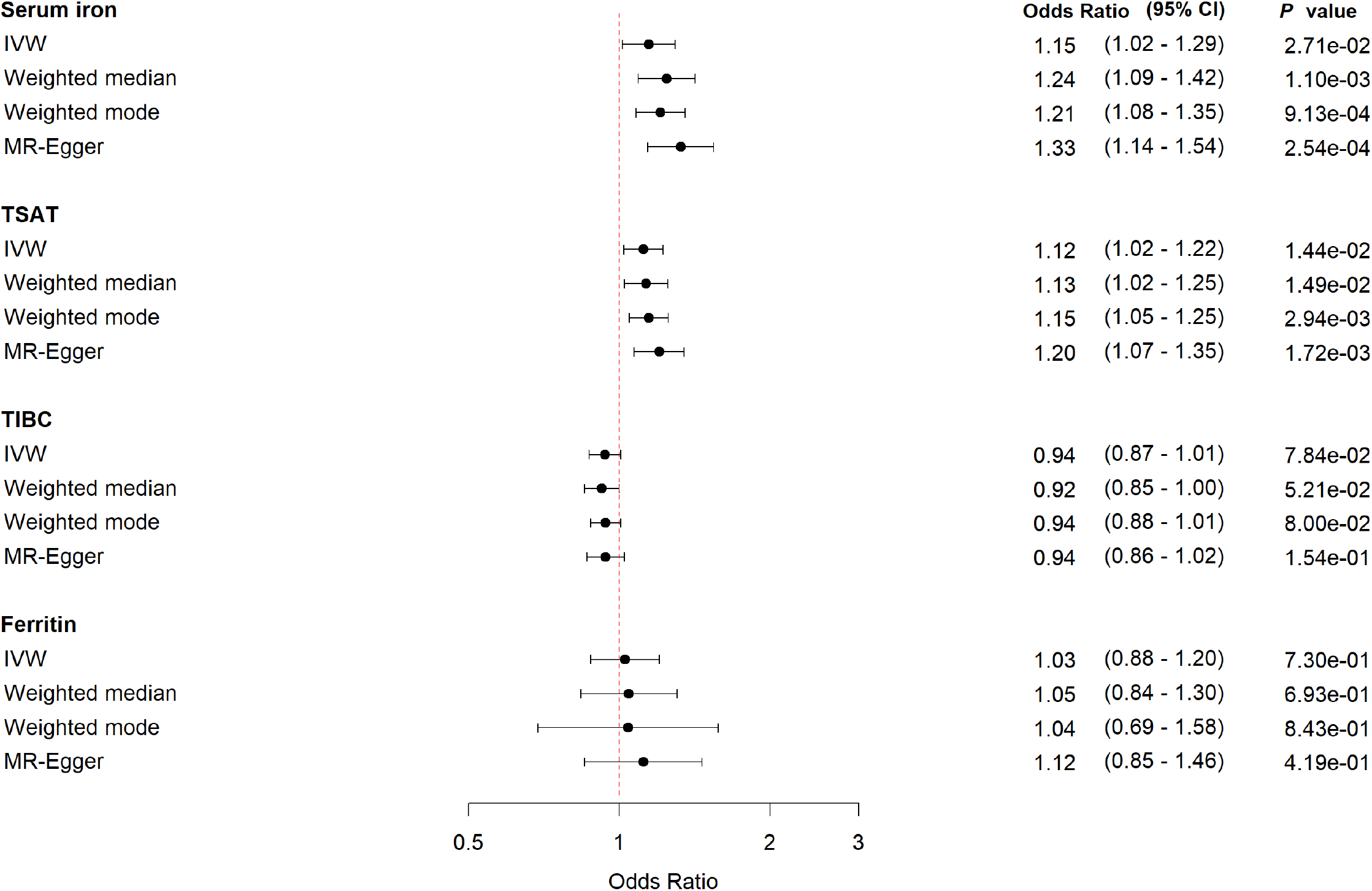
Forest plot with MR estimates for risk of sepsis

Using PhenoScanner, we identified the SNP rs2228145, an instrument for serum iron, to be strongly associated with the IL6-receptor, which we considered a potential biasing pathway due to pleiotopy (Supplemental Table 2). In addition, some of the SNPs used were associated with BMI, CRP, coronary artery disease, triglyceride levels, cholesterol levels, blood pressure, diabetes 2, glycosylated hemoglobin and white blood cell counts. The leave-one-out analyses yielded similar results, suggesting that the different potentially pleiotropic pathways did not substantially affect the results (Supplementary Figure S1).

### COVID-19

We found indication of a relationship between genetically-predicted higher levels of serum iron and risk of being hospitalized with COVID-19 compared with non-hospitalized COVID cases; OR 1.29 (95% CI 0.97 – 1.72, *P*= 0.08) (Figure 2). Similar associations were observed for TSAT and ferritin, but less pronounced. The sensitivity analyses supported the IVW analyses, and leave-one-out plots suggested no pleiotropic effects (Supplemental Figure S2)

**Figure 2:**
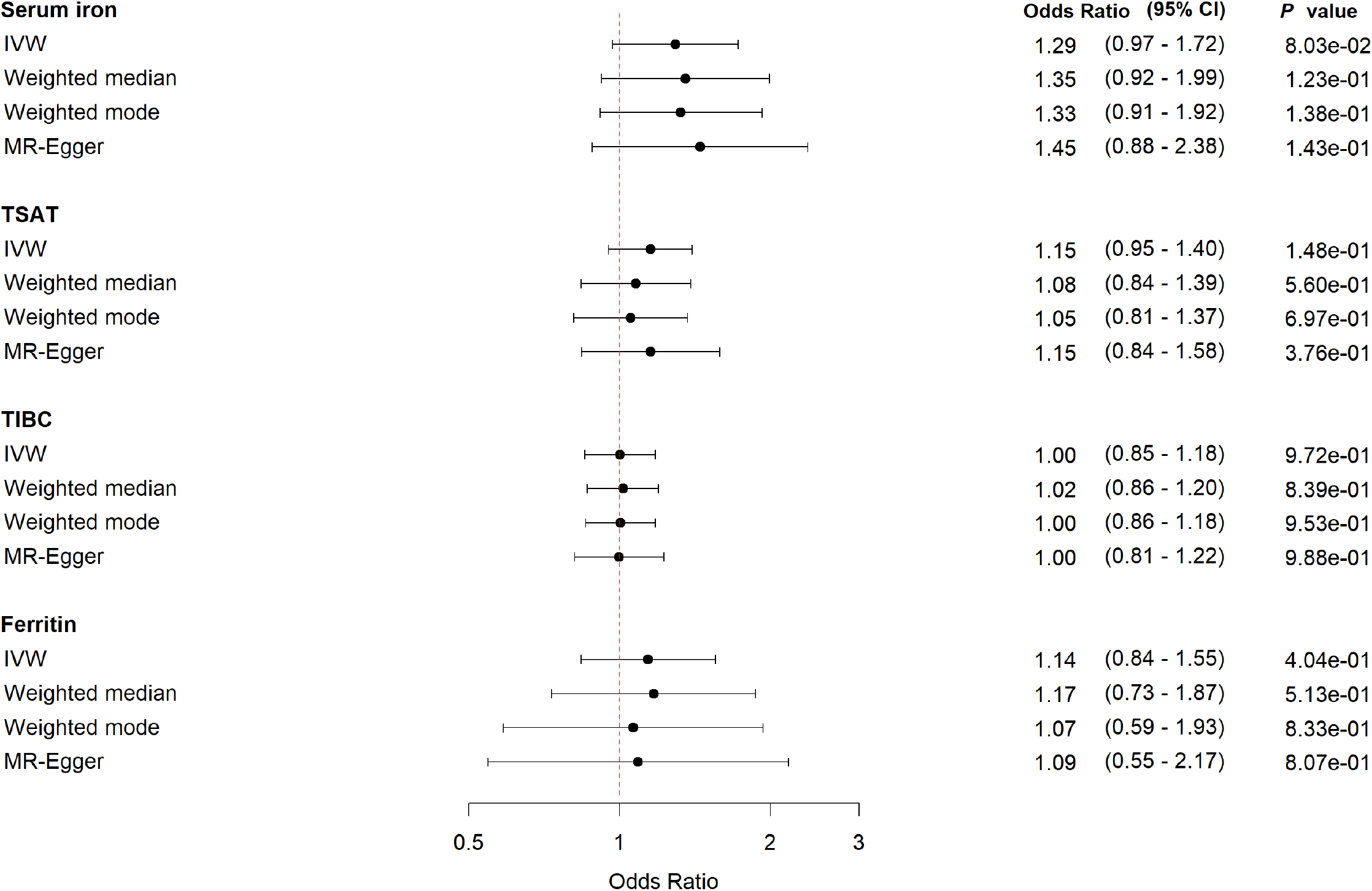
Forest plot with MR estimates for risk of being hospitalized with COVID-19 compared to non-hospitalized COVID-19

In the sex-stratified analyses, we found tendency among women of a harmful effect of increasing genetically-predicted levels of serum iron; OR 1.63 (95% CI 0.94 – 2.86, *P*= 0.09) and TSAT; OR 1.31 (95% CI 0.99 – 1.75, *P*= 0.06). For TIBC and ferritin the estimates were uncertain (Figure 3). The corresponding results for men were less pronounced, and the wide confidence intervals made comparison between the sexes inappropriate (Figure 4) The sensitivity analyses supported the main findings (Figures 3 and 4, and Supplementary Figures S3 and S4).

**Figure 3:**
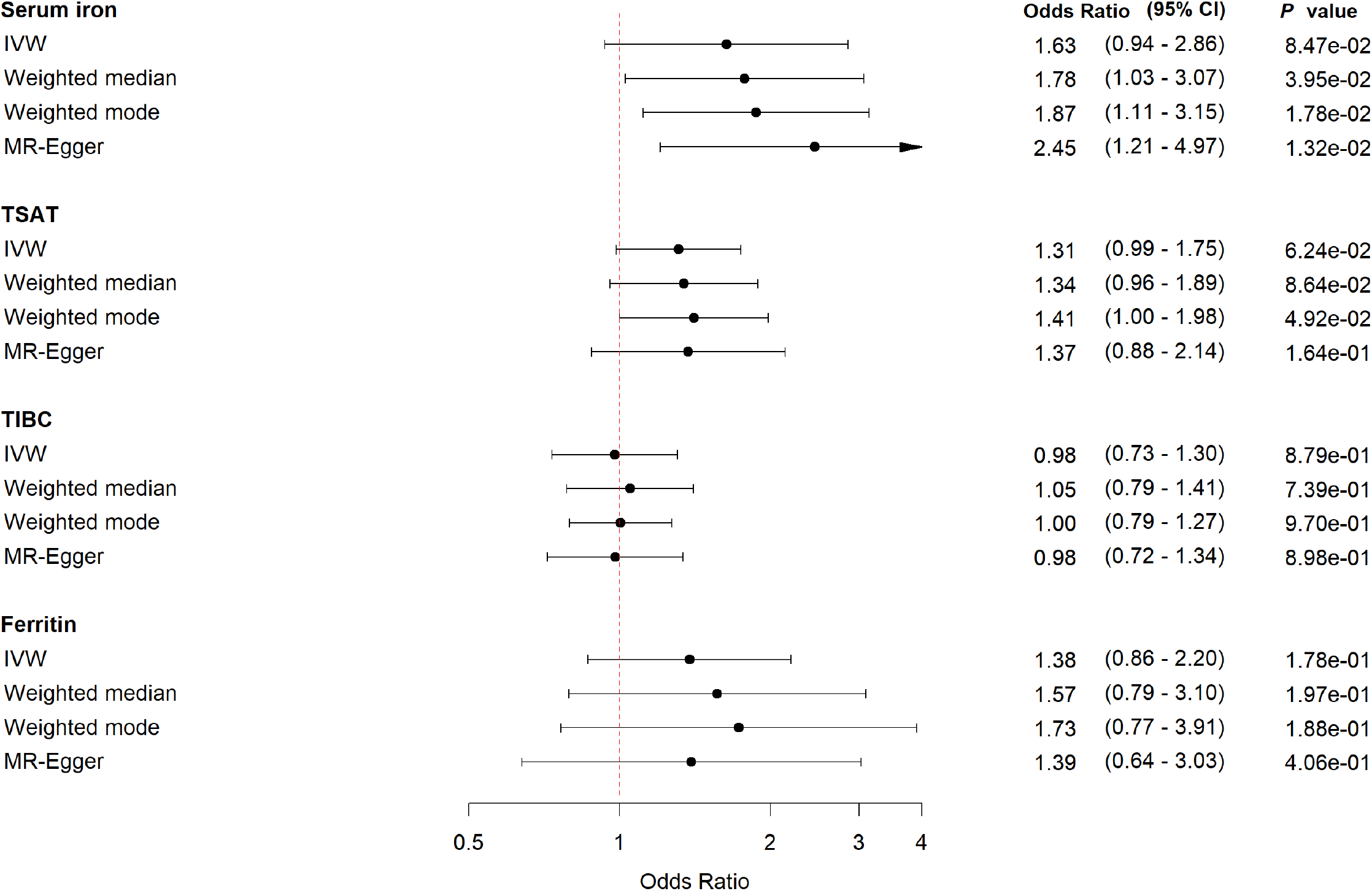
Forest plot for *women* with MR estimates for risk of being hospitalized with COVID-19 compared to non-hospitalized COVID-19

**Figure 4:**
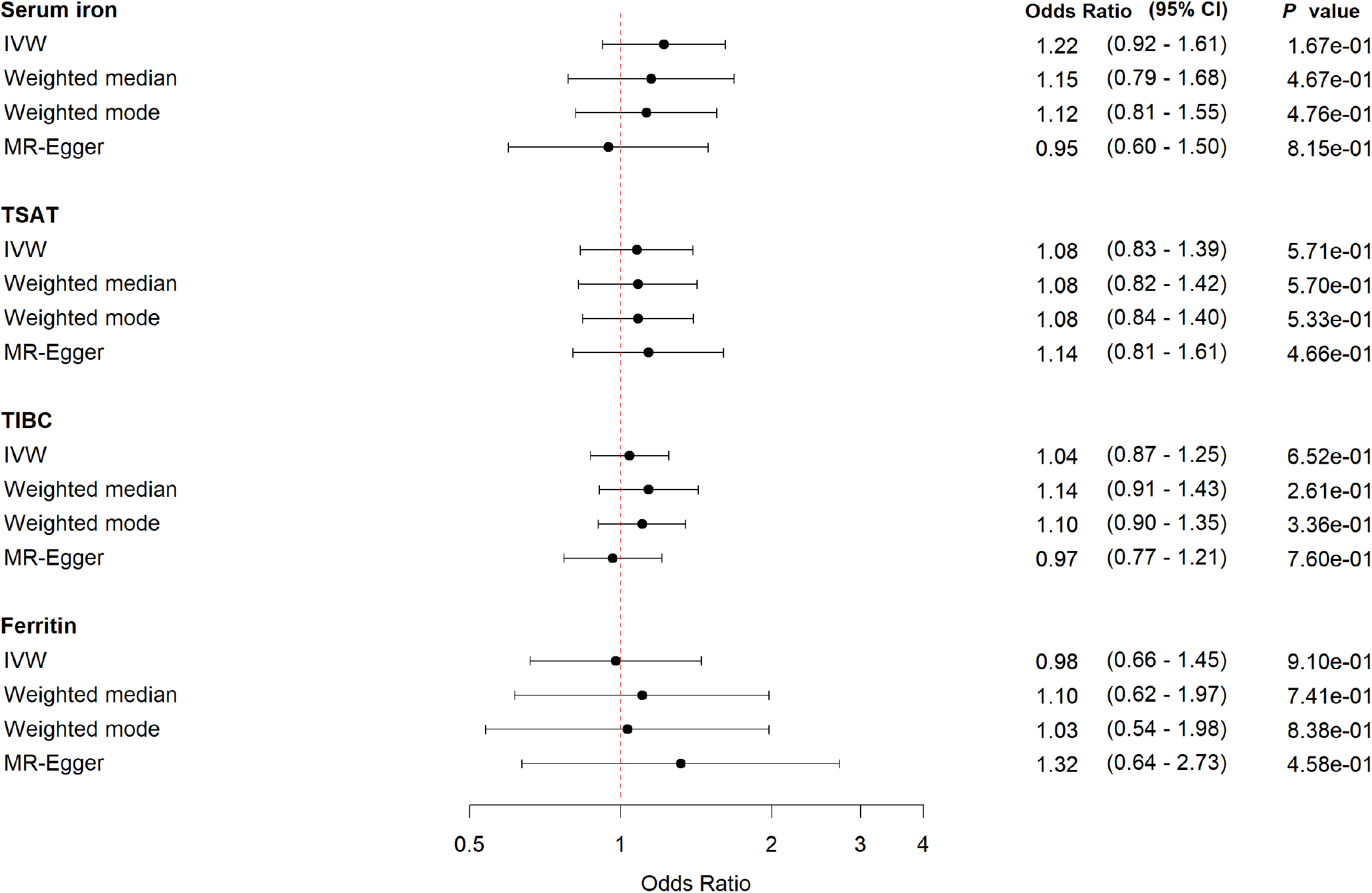
Forest plot for *men* with MR estimates for risk of being hospitalized with COVID-19 compared to non-hospitalized COVID-19

There was no clear evidence that genetically-predicted levels of iron status biomarkers were associated with risk of being hospitalized with COVID-19 compared with the non-hospitalized population including the sex-stratified analyses (Supplemental Figures S5, S6 and S7).

## Discussion

In this study we performed two sample MR analyses to estimate the unconfounded effect of iron status on risk of sepsis and severe COVID-19 using data from large GWASs. The MR results provided some evidence that higher genetically-proxied iron load – reflected in higher levels of serum iron and TSAT, and lower levels of TIBC – were associated with increased risk of sepsis. We found a tendency for increased risk of being hospitalized with COVID-19 compared to non-hospitalized COVID-19 cases in subjects with genetically-predicted higher levels of serum iron. We included sex stratified analyses to assess potential sex differences in the effect of iron status on risk of COVID-19 hospitalizations, which provided some indication of a more pronounced harmful effect of high iron status among women compared with men, but with too little precision to strongly support a difference. The sensitivity analyses supported the overall findings.

Our results were consistent with previous observational studies on sepsis, including a prospective study from Turkey found higher serum iron in septic patients compared to healthy volunteers (8). There is a substantial lack of prospective studies investigating the effect of iron status measured before the onset of the infection. In a prospective population-based cohort study from Norway, we found low iron status to be associated with increased risk of future bloodstream infections (6). This is discordant to our MR results where higher genetically-predicted iron status is related to increased risk of sepsis and being hospitalized due to COVID-19, and could be attributed to differences in the epidemiological methods applied, such as residual confounding, but also limitations with the two-sample MR method used that is restricted to assess linear models (37).

Few MR studies have explored iron status and risk of severe infections. An MR-study using iron related SNPs identified in the Genetics of Iron Status-consortia (38) found evidence that higher serum-iron, TSAT and ferritin were related to increased risk of sepsis (21). Using a more updated set of genetic instruments for iron status biomarkers, we replicated these findings for serum iron and TSAT, a tendency for TIBC, but not for ferritin. Another MR study found evidence of increased risk of skin and soft-tissue infections with higher serum iron levels (39).

To the best of our knowledge, no previous study has conducted MR analysis to investigate the effect of iron status on incidence or outcome of COVID-19. Observational studies that have investigated iron status at the time of infection and found evidence of low iron status being a risk factor for a severe course of COVID-19 (12). Another study with COVID-19 patients compared to non-COVID-19 patients showed lower serum iron and TSAT levels in patients with COVID-19 independently of severity. Whereas COVID-19 patients defined as severe and critical had substantially higher ferritin levels (40). Some have linked COVID-19 to the hyperferritinemic syndromes which is associated with hyperinflammation (41). We identified a strong tendency towards an increased risk of being hospitalized with COVID-19 in persons with genetically proxied higher iron status. Differences between our findings and those reported in observational studies could reflect the fact that associations between iron status and COVID-19 may be confounded by factors difficult to adjust for such as poor nutritional status (4) or medical comorbidities associated with functional iron deficiency (42). Despite numerous observational studies in COVID-19 patients, the role of iron status before the time of infection as well as changes in iron status during infection has not been ruled out and the same applies to sepsis. We hypothesize that individuals with higher iron status could be less able to handle the acute iron load seen during severe infections, leaving them more vulnerable to be hospitalized with sepsis and COVID-19.

The role of iron status in the context of infectious diseases has long been noted (1, 2, 10, 11). Both iron deficiency (5, 6), iron overload (8), and iron fortification programs without adequate infection surveillance (43), have been linked to increased risk of infections. To date, treatment with iron chelators in sepsis or COVID-19 have not been studied in any large RCT, although suggested as potential adjuvant therapy in several reviews (44, 45). Experimental models of sepsis studying different iron chelators, report promising anti-inflammatory and anti-bacterial effects (46). One pilot study in 92 COVID-19 patients using oral and intranasal lactoferrin have shown promising results with faster clinical symptoms recovery and lower serum ferritin levels in patients with mild to moderate COVID-19 compared to controls (47).

The pathway from iron status to risk of severe infections like sepsis and COVID-19 could be multifactorial, including long-term effects of iron status on immune functions and susceptibility to pathogens, but also adaptations in iron status at the time of infection (2, 11, 44). We identified that iron status affects the risk of sepsis and the risk of being hospitalized with COVID-19, indicating that iron status before the time of infection interfere with the response to infection.

It is well established that iron status varies according to sex (3, 4). Observational studies have shown that men are more prone to a severe course of COVID-19 (18, 19), leaving sex-stratified investigations important to reveal potential explanations for the sex differences (23). We assessed sex specific summary-level data on iron status and COVID-19 outcomes and showed that there was some tendency that the effect of serum iron and TSAT was more pronounced among women compared with men. Another study looking at iron status at time of hospitalization for COVID-19 identified sex differences where female patients had significantly lower serum iron, TSAT and ferritin levels and higher TIBC levels compared to men, whereas the association with severity between serum iron and TSAT was observed in both sexes (15).

Major strengths with our study include the use of large GWAS summary data for both iron status, sepsis, and severe COVID-19. Our main MR estimates were similar using IVW, weighted median, weighted mode, and MR Egger methods. As MR studies could carry the risk of pleiotropy, we used various strategies to detect and account for the potential pleiotropy. Taken together, the overall conclusions of our study were less likely to be affected by bias due to pleiotropy. We used GWAS summary data from European decent to reduce confounding due to population stratification. The slight difference in estimation and confidence intervals between the different MR methods were expected and most likely do not represent actual differences (48).

Several limitations should be considered in our study. First, the participants in our study are restricted to European ancestry and as both severe infections and iron deficiency are global concerns, our findings should be examined in other populations. Second, the sepsis phenotype has proven to be heterogeneous depending on how the causal pathogen act on the host immune functions and factors within the host (49). Timing and correct treatment of infections before they evolve to sepsis, further access to organ supportive treatment in intensive care units, and severity of sepsis might also be different. During the COVID-19 pandemic limited hospital resources and capacity might have influenced on hospitalizations. Third, iron status changes substantially during infection and inflammation, further exacerbated by tissue destruction and cell death. Iron status fluctuates during a lifetime, during periods of higher demand and need such as pregnancy and growth, in situations with increased losses (i.e. blood loss or critical illness), and due to chronical medical disorders (50). Genetically-predicted iron status may not perfectly reflect this time-varying exposure (51). The U-shaped risk relationship that has been proposed for the extremes of iron status (10, 15) might cause an attenuated association when evaluated in a linear model as in the two-sample MR methods. Non-linear MR methods could be more suitable to explore this U-shaped relationship but requires large GWAS with both measurements of iron biomarkers as well as the outcomes of interest (37). However, observational studies measuring iron status at time of infection might be biased by the acute phase response leading to iron depletion and hyperferritinemia (i.e. reverse causation) which we avoided using an MR framework. Due to the MR methods’ use of genetic instruments, the possibility of confounding was limited. Using sex stratified summary-level data for both exposure and COVID-19 outcomes we were able to investigate potential sex differences in the associations.

In conclusion, our study leveraged large-scale summary data to explore the effects of iron status on risk of sepsis and severe COVID-19. Our findings support a causal association between high iron status and increased risk of sepsis and in verified cases of COVID-19 we identified a tendency of higher risk of being hospitalized in persons with higher iron status. We highlight the importance of sex specified summary-level data to assess potential sex differences in the associations. For being hospitalized with COVID-19 there were indications of a more pronounced effect of higher iron status in women compared to men. Future studies are needed to explore the exact mechanisms of iron status and severe infections with the potential of prevention management and treatment strategies.

## Data Availability

All data are publicly available as summary-level data from: gwas.mrcieu.ac.uk/, covid19hg.org, grasp.nhlbi.nih.gov/covid19GWASResults.aspx

http://covid19hg.org

http://grasp.nhlbi.nih.gov/covid19GWASResults.aspx

## Consent for publication

No individually identifiable data was included in this manuscript.

## Authors’ contributions

RMM and TR originated the research hypothesis and conceptual design. RMM and HMF analyzed the data and prepared tables and figures. RMM drafted the manuscript. HMF, KVL, ATD, ES, JKD, BOÅ, LTG and TR revised the manuscript. BOÅ and TR provided critical inputs in study methods and analyses. JKD and TR supervised the study. All authors revised and approved the submission.

## Web links

Sepsis GWAS. gwas.mrcieu.ac.uk/

COVID-19 Host Genetics Initiative. covid19hg.org

Sex disaggregated COVID-19 GWAS. NHLBI GRASP catalogue, release date 06.18.21, grasp.nhlbi.nih.gov/covid19GWASResults.aspx

## References

1. Beard JL. Iron biology in immune function, muscle metabolism and neuronal functioning. J Nutr. 2001;131(2s-2):568S–79S; discussion 80S.

2. Ganz T, Nemeth E. Iron homeostasis in host defence and inflammation. Nature reviews Immunology. 2015;15(8):500–10.

3. WHO. WHO guideline on use of ferritin concentrations to assess iron status in individuals and populations. Geneva: World Health Organization; 2020. p. 72.

4. WHO. Assessing the iron status of populations: including litterature reviews. Geneva: World Health Organization; 2007. p. 108.

5. Tansarli GS, Karageorgopoulos DE, Kapaskelis A, Gkegkes I, Falagas ME. Iron deficiency and susceptibility to infections: evaluation of the clinical evidence. Eur J Clin Microbiol Infect Dis. 2013;32(10):1253–8.

6. Mohus RM, Paulsen J, Gustad L, Askim Å, Mehl A, DeWan AT, et al. Association of iron status with the risk of bloodstream infections: results from the prospective population-based HUNT Study in Norway. Intensive Care Med. 2018;44(8):1276–83.

7. Brandtner A, Tymoszuk P, Nairz M, Lehner GF, Fritsche G, Vales A, et al. Linkage of alterations in systemic iron homeostasis to patients’ outcome in sepsis: a prospective study. J Intensive Care. 2020;8:76.

8. Akkas I, Ince N, Sungur MA. Serum trace element and heavy metal levels in patients with sepsis. Aging Male. 2020;23(3):222–6.

9. Lan P, Pan K-h, Wang S-j, Shi Q-c, Yu Y-x, Fu Y, et al. High Serum Iron level is Associated with Increased Mortality in Patients with Sepsis. Scientific Reports. 2018;8(1):11072.

10. Swenson ER, Porcher R, Piagnerelli M. Iron deficiency and infection: another pathway to explore in critically ill patients? Intensive Care Medicine. 2018;44(12):2260–2.

11. Drakesmith H, Prentice AM. Hepcidin and the Iron-Infection Axis. Science (New York, NY). 2012;338(6108):768–72.

12. Lv Y, Chen L, Liang X, Liu X, Gao M, Wang Q, et al. Association between iron status and the risk of adverse outcomes in COVID-19. Clin Nutr. 2021;40(5):3462–9.

13. Hippchen T, Altamura S, Muckenthaler MU, Merle U. Hypoferremia predicts hospitalization and oxygen demand in COVID-19 patients. medRxiv. 2020:2020.06.26.20140525.

14. Perricone C, Bartoloni E, Bursi R, Cafaro G, Guidelli GM, Shoenfeld Y, et al. COVID-19 as part of the hyperferritinemic syndromes: the role of iron depletion therapy. Immunol Res. 2020;68(4):213–24.

15. Tojo K, Sugawara Y, Oi Y, Ogawa F, Higurashi T, Yoshimura Y, et al. The U-shaped association of serum iron level with disease severity in adult hospitalized patients with COVID-19. Scientific Reports. 2021;11(1):13431.

16. Galmés S, Serra F, Palou A. Current State of Evidence: Influence of Nutritional and Nutrigenetic Factors on Immunity in the COVID-19 Pandemic Framework. Nutrients. 2020;12(9).

17. Mohus RM, Gustad LT, Furberg AS, Moen MK, Liyanarachi KV, Askim Å, et al. Explaining sex differences in risk of bloodstream infections using mediation analysis in the population-based HUNT study in Norway. Sci Rep. 2022;12(1):8436.

18. Alwani M, Yassin A, Al-Zoubi RM, Aboumarzouk OM, Nettleship J, Kelly D, et al. Sex-based differences in severity and mortality in COVID-19. Reviews in Medical Virology. 2021;31(6):e2223.

19. Peckham H, de Gruijter NM, Raine C, Radziszewska A, Ciurtin C, Wedderburn LR, et al. Male sex identified by global COVID-19 meta-analysis as a risk factor for death and ITU admission. Nature Communications. 2020;11(1):6317.

20. Davies NM, Holmes MV, Davey Smith G. Reading Mendelian randomisation studies: a guide, glossary, and checklist for clinicians. Bmj. 2018;362:k601.

21. Hu Y, Cheng X, Mao H, Chen X, Cui Y, Qiu Z. Causal Effects of Genetically Predicted Iron Status on Sepsis: A Two-Sample Bidirectional Mendelian Randomization Study. Front Nutr. 2021;8:747547.

22. Bell S, Rigas AS, Magnusson MK, Ferkingstad E, Allara E, Bjornsdottir G, et al. A genome-wide meta-analysis yields 46 new loci associating with biomarkers of iron homeostasis. Commun Biol. 2021;4(1):156.

23. Mauvais-Jarvis F, Bairey Merz N, Barnes PJ, Brinton RD, Carrero JJ, DeMeo DL, et al. Sex and gender: modifiers of health, disease, and medicine. Lancet. 2020;396(10250):565–82.

24. Burgess S, Thompson SG. Avoiding bias from weak instruments in Mendelian randomization studies. International journal of epidemiology. 2011;40(3):755–64.

25. Hemani G, Zheng J, Elsworth B, Wade KH, Haberland V, Baird D, et al. The MR-Base platform supports systematic causal inference across the human phenome. Elife. 2018;7.

26. Broadbent JR, Foley CN, Grant AJ, Mason AM, Staley JR, Burgess S. MendelianRandomization v0.5.0: updates to an R package for performing Mendelian randomization analyses using summarized data. Wellcome Open Res. 2020;5:252.

27. Rudd KE, Johnson SC, Agesa KM, Shackelford KA, Tsoi D, Kievlan DR, et al. Global, regional, and national sepsis incidence and mortality, 1990-2017: analysis for the Global Burden of Disease Study. Lancet. 2020;395(10219):200–11.

28. Elsworth B, Lyon M, Alexander T, Liu Y, Matthews P, Hallett J, et al. The MRC IEU OpenGWAS data infrastructure. bioRxiv. 2020:2020.08.10.244293.

29. Mapping the human genetic architecture of COVID-19. Nature. 2021;600(7889):472–7.

30. Leslie R, O’Donnell CJ, Johnson AD. GRASP: analysis of genotype-phenotype results from 1390 genome-wide association studies and corresponding open access database. Bioinformatics (Oxford, England). 2014;30(12):i185–i94.

31. Burgess S, Davey Smith G, Davies NM, Dudbridge F, Gill D, Glymour MM, et al. Guidelines for performing Mendelian randomization investigations. Wellcome Open Res. 2019;4:186.

32. Bowden J, Davey Smith G, Haycock PC, Burgess S. Consistent Estimation in Mendelian Randomization with Some Invalid Instruments Using a Weighted Median Estimator. Genet Epidemiol. 2016;40(4):304–14.

33. Hartwig FP, Davey Smith G, Bowden J. Robust inference in summary data Mendelian randomization via the zero modal pleiotropy assumption. International journal of epidemiology. 2017;46(6):1985–98.

34. Bowden J, Davey Smith G, Burgess S. Mendelian randomization with invalid instruments: effect estimation and bias detection through Egger regression. International journal of epidemiology. 2015;44(2):512–25.

35. Kamat MA, Blackshaw JA, Young R, Surendran P, Burgess S, Danesh J, et al. PhenoScanner V2: an expanded tool for searching human genotype-phenotype associations. Bioinformatics. 2019;35(22):4851–3.

36. Skrivankova VW, Richmond RC, Woolf BAR, Davies NM, Swanson SA, VanderWeele TJ, et al. Strengthening the reporting of observational studies in epidemiology using mendelian randomisation (STROBE-MR): explanation and elaboration. BMJ. 2021;375:n2233.

37. Burgess S, Davies NM, Thompson SG. Instrumental variable analysis with a nonlinear exposure-outcome relationship. Epidemiology. 2014;25(6):877–85.

38. Benyamin B, Esko T, Ried JS, Radhakrishnan A, Vermeulen SH, Traglia M, et al. Novel loci affecting iron homeostasis and their effects in individuals at risk for hemochromatosis. Nat Commun. 2014;5:4926.

39. Gill D, Benyamin B, Moore LSP, Monori G, Zhou A, Koskeridis F, et al. Associations of genetically determined iron status across the phenome: A mendelian randomization study. PLoS Med. 2019;16(6):e1002833.

40. Bastin A, Shiri H, Zanganeh S, Fooladi S, Momeni Moghaddam MA, Mehrabani M, et al. Iron Chelator or Iron Supplement Consumption in COVID-19? The Role of Iron with Severity Infection. Biol Trace Elem Res. 2021:1–11.

41. Colafrancesco S, Alessandri C, Conti F, Priori R. COVID-19 gone bad: A new character in the spectrum of the hyperferritinemic syndrome? Autoimmun Rev. 2020;19(7):102573-.

42. Weiss G, Goodnough LT. Anemia of chronic disease. N Engl J Med. 2005;352(10):1011–23.

43. Sazawal S, Black RE, Ramsan M, Chwaya HM, Stoltzfus RJ, Dutta A, et al. Effects of routine prophylactic supplementation with iron and folic acid on admission to hospital and mortality in preschool children in a high malaria transmission setting: community-based, randomised, placebo-controlled trial. Lancet. 2006;367(9505):133–43.

44. Habib HM, Ibrahim S, Zaim A, Ibrahim WH. The role of iron in the pathogenesis of COVID-19 and possible treatment with lactoferrin and other iron chelators. Biomedicine & Pharmacotherapy. 2021;136:111228.

45. Poonkuzhi Naseef P, Elayadeth-Meethal M, Mohammed Salim KT, Anjana A, Muhas C, Abdul Vajid K, et al. Therapeutic potential of induced iron depletion using iron chelators in Covid-19. Saudi Journal of Biological Sciences. 2022;29(4):1947–56.

46. Lehmann C, Aali M, Zhou J, Holbein B. Comparison of Treatment Effects of Different Iron Chelators in Experimental Models of Sepsis. Life (Basel). 2021;11(1):57.

47. Campione E, Lanna C, Cosio T, Rosa L, Conte MP, Iacovelli F, et al. Lactoferrin as potential supplementary nutraceutical agent in COVID-19 patients: <em>in vitro</em> and <em>in vivo</em> preliminary evidences. bioRxiv. 2020:2020.08.11.244996.

48. Slob EAW, Burgess S. A comparison of robust Mendelian randomization methods using summary data. Genet Epidemiol. 2020;44(4):313–29.

49. Leligdowicz A, Matthay MA. Heterogeneity in sepsis: new biological evidence with clinical applications. Critical care (London, England). 2019;23(1):80-.

50. WHO. Assessing the iron status of populations: including literature reviews. Geneva: World Health Organization; 2007.

51. Labrecque JA, Swanson SA. Interpretation and Potential Biases of Mendelian Randomization Estimates With Time-Varying Exposures. Am J Epidemiol. 2019;188(1):231–8.

