## Supplemental material for "Iron status and the risk of sepsis and severe COVID-19: A two-sample Mendelian randomization study"

Helene Flatby^1^

Kristin V. Liyanarachi^1,3^

Andrew T. DeWan^4,1^

Erik Solligård^1^

Jan Kristian Damås^1,3,5^

Bjørn Olav Åsvold^6,7^

Lise T. Gustad^1,8,9^

Tormod Rogne^4,1,10^

**Supplemental Table S1: Included SNPs for all and sex-disaggregated analyses with explained variance and F-statistics for each iron biomarker**

|  |  | **All** | | | | **Women** | | | | **Men** | | | |
| --- | --- | --- | --- | --- | --- | --- | --- | --- | --- | --- | --- | --- | --- |
|  |  | Serum iron | TSAT | TIBC | Ferritin | Serum iron | TSAT | TIBC | Ferritin | Serum iron | TSAT | TIBC | Ferritin |
| No of SNPs | Sepsis | 14 | 10 | 14 | 33 | NA | NA | NA | NA | NA | NA | NA | NA |
|  | Covid-19 | 11 | 9 | 11 | 27 | 14 | 9 | 15 | 33 | 14 | 9 | 15 | 33 |
| Median variance explained (%) | | 1.8 % | 3.4 % | 2.2 % | 1.5 % | 2.0% | 3.5% | 1.9 % | 1.5% | 1.9% | 3.5% | 2.0% | 1.4% |
| Range Variance explained (%) | | 1.4– 7.5 % | 1.7– 9.7 % | 1.4–12.0% | 1.1– 3.7% | 1.1– 9.4% | 1.4– 10.5% | 1.3– 10.3% | 0.6– 4.9% | 0.9– 9.8% | 1.3– 12.5% | 0.5– 12.3% | 0.7– 3.8% |
| Median F-statistic | | 291 | 550 | 233 | 158 | 148 | 334 | 138 | 61 | 96 | 232 | 101 | 44 |
| Range F-statistic | | 226–1288 | 286– 1680 | 147– 1413 | 115– 397 | 79– 766 | 127–1076 | 90– 814 | 22–205 | 49– 543 | 82–924 | 27–702 | 22–122 |

TSAT; transferrin saturation, TIBC; total iron binding capacity, SNP; single nucleotide polymorphism, NA; not available

**Supplemental Table S2: Phenoscanner results for included SNPs linked to other biological traits than iron status**

| **SNP** | **Trait** | **BETA** | ***P* value** |
| --- | --- | --- | --- |
| rs1250259 | Total cholesterol | 0.03 | 7.56e-06 |
| rs1250259 | LDL cholesterol | 0.03 | 1.46e-06 |
| rs1250259 | Blood pressure | -0.02 | 1.57e-11 |
| rs1260326 | Triglyserides | -0.12 | 4.00e-253 |
| rs1260326 | Total cholesterol | -0.05 | 3.00e-42 |
| rs1260326 | Diabetes 2 | 0.08 | 3.70e-09 |
| rs1260326 | Neutrophil count | -0.03 | 1.16e-19 |
| rs1260326 | Lymphocyte count | 0.03 | 2.00e-12 |
| rs1260326 | CRP | -0.07 | 5.00e-40 |
| rs12807014 | BMI | -0.02 | 1.49e-15 |
| rs2954029 | Triglycerides | -0.08 | 1.00e-107 |
| rs2954029 | Total cholesterol | 0.06 | 1.00e-77 |
| rs2954029 | Coronary artery disease | 0.05 | 1.68e-22 |
| rs2954029 | Granulocyte count | -0.02 | 5.08e-10 |
| rs2954029 | Neutrophil count | -0.03 | 8.13e-12 |
| rs2954029 | Lymphocyte count | 0.03 | 1.17e-13 |
| rs2954029 | BMI | NA | NA |
| rs34523089 | Monocyte count | -0.05 | 1.77e-21 |
| rs34523089 | Granulocyte count | 0.05 | 1.71e-26 |
| rs3743171 | Monocyte count | 0.04 | 6.31e-14 |
| rs3743171 | Granulocyte count | -0.04 | 7.13e-14 |
| rs3743171 | Neutrophile count | -0.02 | 1.96e-06 |
| rs3743171 | BMI | 0.01 | 6.50e-06 |
| rs4808802 | Total cholesterol | 0.03 | 3.27e-08 |
| rs4808802 | Granulocyte count | 0.02 | 4.72e-06 |
| rs55789050 | Diabetes 2 | -0.06 | 7.80e-07 |
| rs601338 | Total cholesterol | -0.03 | 2.41e-10 |
| rs174546 | Neutrophile count | 0.02 | 3.11e-10 |
| rs174546 | Monocyte count | -0.03 | 2.71e-14 |
| rs174546 | Granulocyte count | 0.03 | 5.03e-16 |
| rs174546 | Triglycerides | -3.82 | 5.00e-24 |
| rs174546 | Total cholesterol | 0.05 | 2.67e-37 |
| rs2228145 | IL-6 | NA | 2.00e-57 |
| rs2228145 | Coronary artery disease | 0.04 | 4.80e-14 |
| rs2228145 | CRP | 0.11 | 1.96e-10 |
| rs2228145 | Granulocyte count | 0.02 | 4.23e-07 |
| rs2228145 | Monocyte count | -0.02 | 8.94e-06 |
| rs35945185 | Lymphocyte count | -0.02 | 2.47e-06 |
| rs35945185 | Granulocyte count | -0.05 | 2.74e-36 |
| rs35945185 | Neutrophile count | -0.05 | 6.22e-36 |
| rs1799945 | Hypertension | -0.1 | 2.00e-10 |
| rs1799945 | HbA1c | 0.02 | 3.76e-19 |
| rs1800562 | HbA1c | -0.04 | 4.67e-28 |
| rs1800562 | Total cholesterol | -0.06 | 1.91e-12 |
| rs855791 | HbA1c | -0.02 | 3.44e-28 |
| rs17580 | Granulocyte count | -0.04 | 6.30e-06 |
| rs59950280 | Triglycerides | 0.04 | 1.00e-10 |
| rs59950280 | Coronary artery disease | 0.04 | 1.00e-06 |
| rs59950280 | Total cholesterol | 0.04 | 1.00e-10 |
| rs9399136 | White blood cell count | -0.05 | 1.65e-29 |

LDL; low density lipoprotein, CRP; C-reactive protein, BMI; body mass index, IL-6; Interleukin-6, HbA1c; glycosylated hemoglobin

**Supplemental Figure S1 (A-D): Leave-one-out plots for the association between the iron biomarkers and sepsis**

A) Serum iron - sepsis

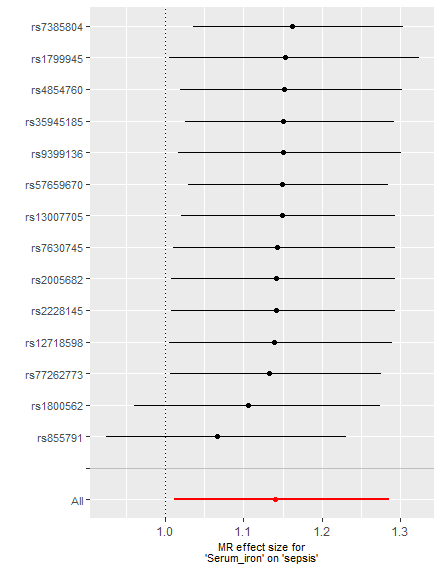

**Odds ratios with 95% CIs**

B) TSAT – sepsis

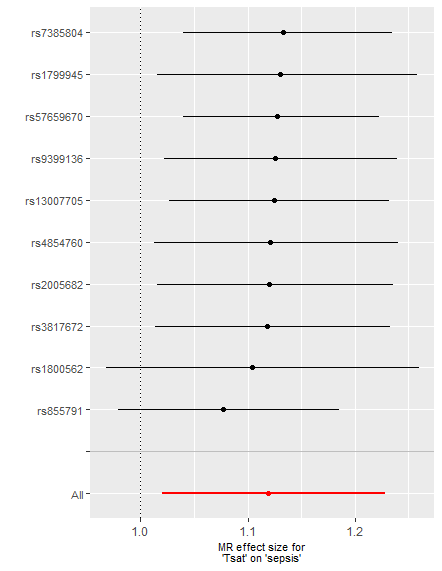

**Odds ratios with 95% CIs**

C) TIBC – sepsis

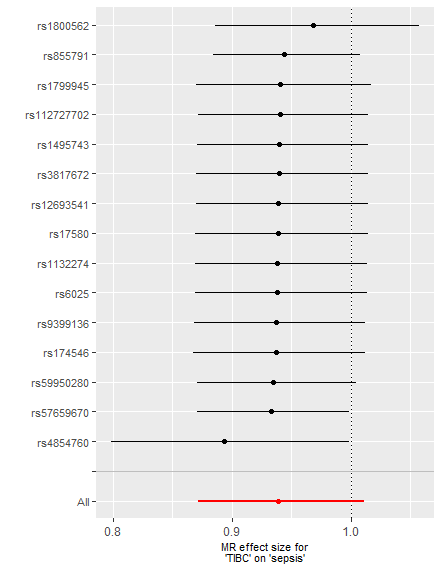

**Odds ratios with 95% CIs**

D) Ferritin – sepsis

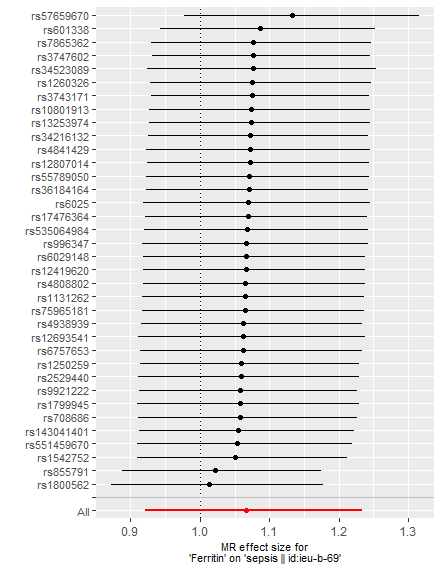

**Odds ratios with 95% CIs**

**Supplemental Figure S2 (A-D): Leave-one-out plots for the association between the iron biomarkers and hospitalized COVID-19 vs non-hospitalized COVID-19**

A) Serum iron – hospitalized COVID-19

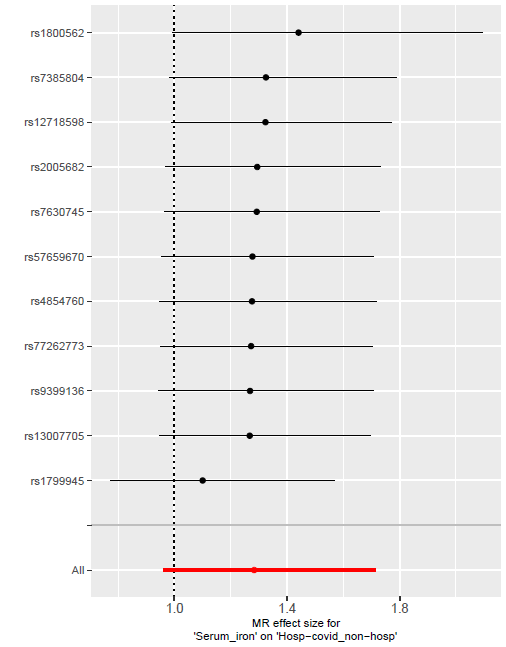

**Odds ratios with 95% Cis**

B) TSAT – hospitalized COVID-19

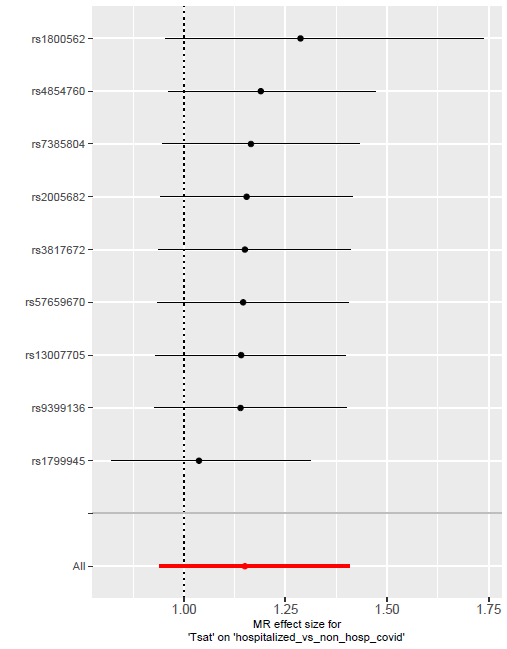

**Odds ratios with 95% CIs**

C) TIBC – hospitalized COVID-19

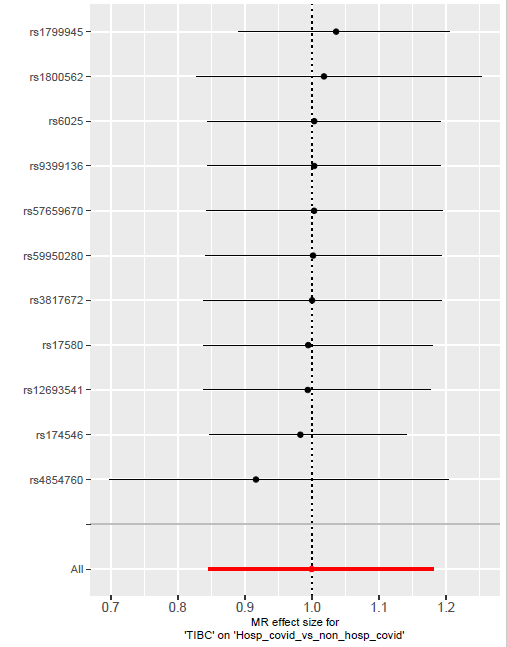

**Odds ratios with 95% CIs**

D) Ferritin – hospitalized COVID-19

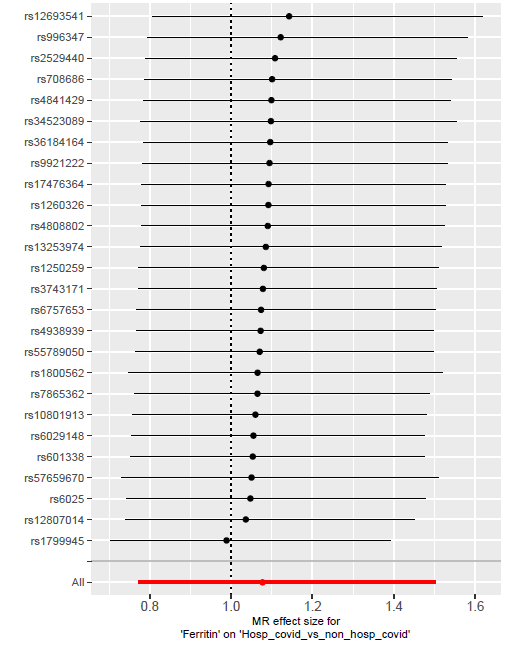

**Odds ratios with 95% CIs**

**Supplemental Figure S3 (A-D): *Women* - Leave-one-out plots for the association between the iron biomarkers and hospitalized COVID-19 vs non-hospitalized COVID-19**

A) Serum iron – hospitalized COVID-19 Women

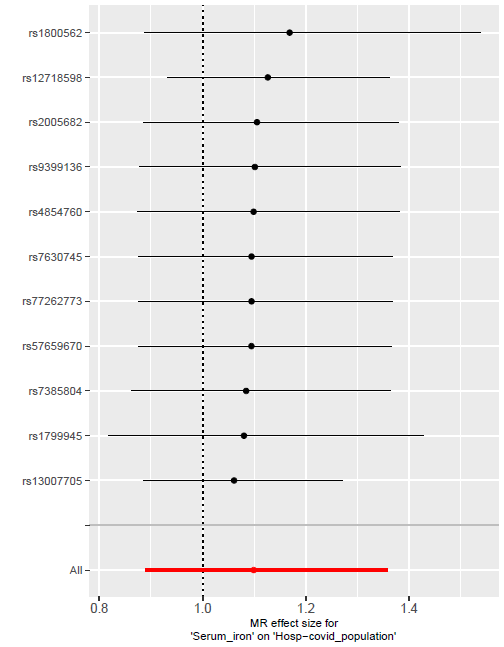

**Odds ratios with 95% CIs**

B) TSAT – hospitalized COVID-19 Women

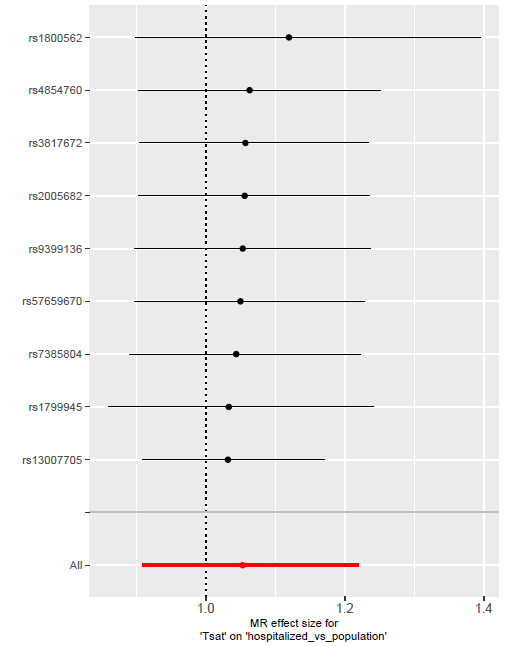

**Odds ratios with 95% CIs**

C) TIBC – hospitalized COVID-19 Women

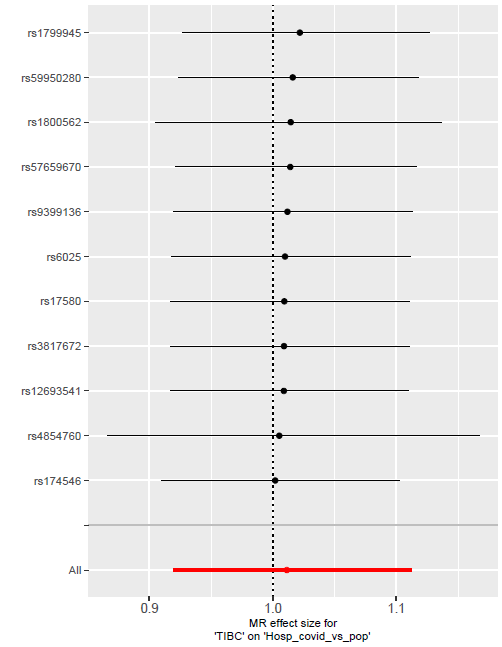

**Odds ratios with 95% Cis**

D) Ferritin – hospitalized COVID-19 Women

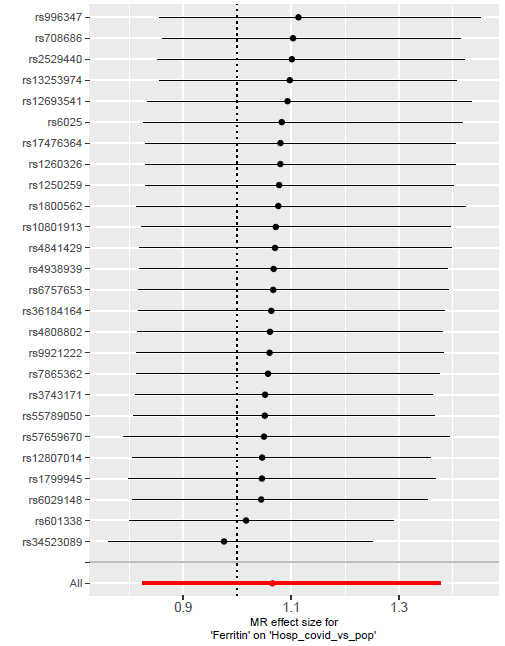

**Odds ratios with 95% CIs**

**Supplemental Figure S3 (A-D): *Men* - Leave-one-out plots for the association between the iron biomarkers and hospitalized COVID-19 vs non-hospitalized COVID-19**

A) Serum iron – hospitalized COVID-19 Men

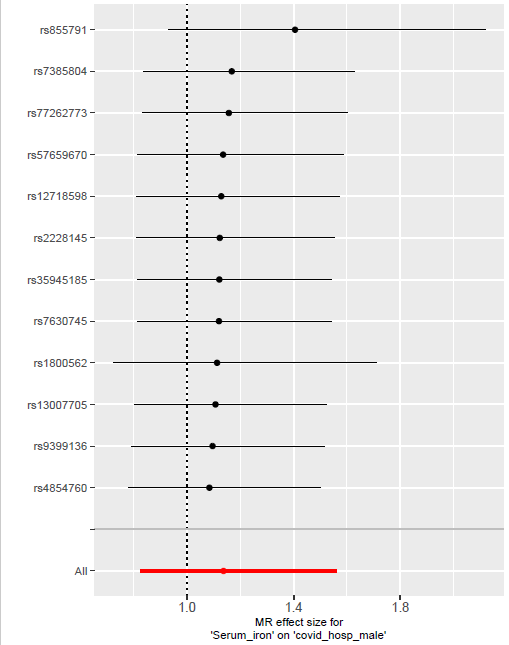

**Odds ratios with 95% Cis**

B) TSAT – hospitalized COVID-19 Men

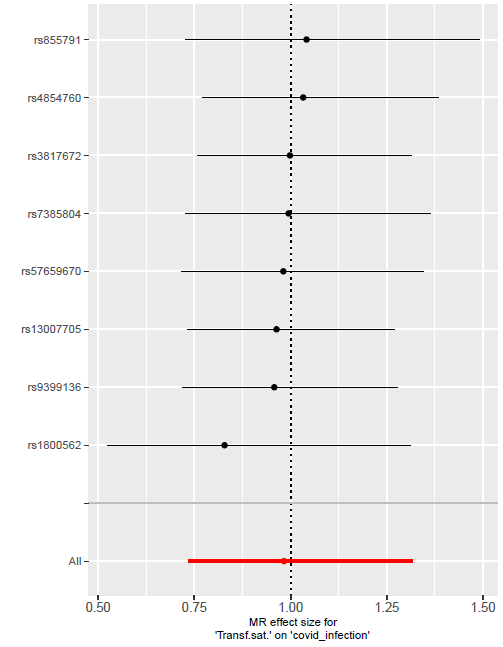

**Odds ratios with 95% CIs**

C) TIBC – hospitalized COVID-19 Men

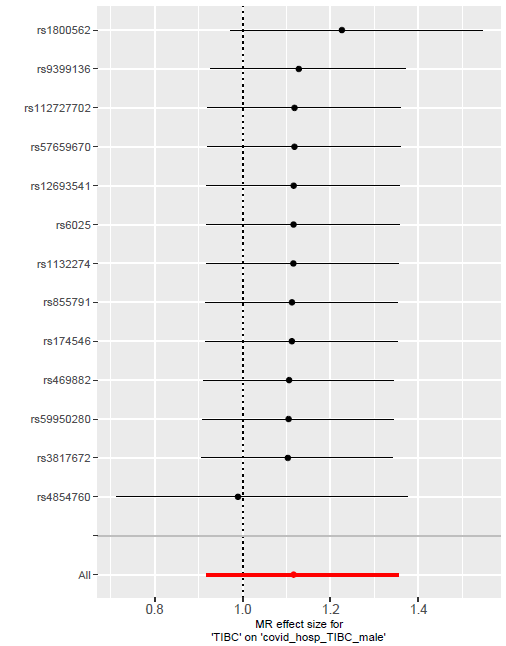

**Odds ratios with 95% CIs**

D) Ferritin – hospitalized COVID-19 Men

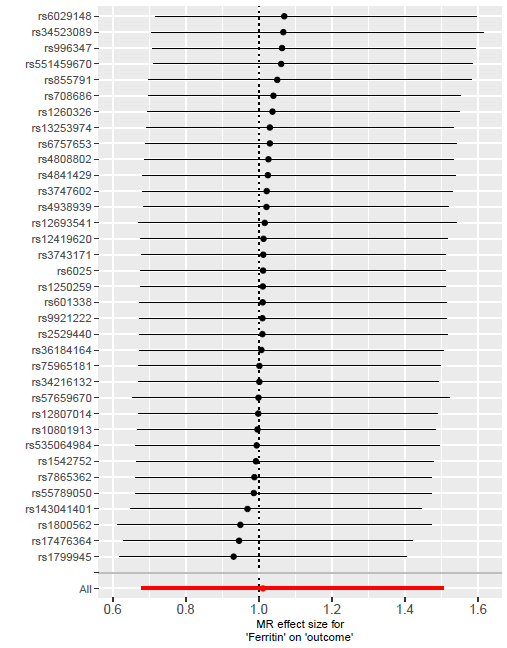

**Odds ratios with 95% CIs**

**Supplemental Figure S5: Forest plot with MR estimates for risk of being hospitalized with COVID-19 compared with population**

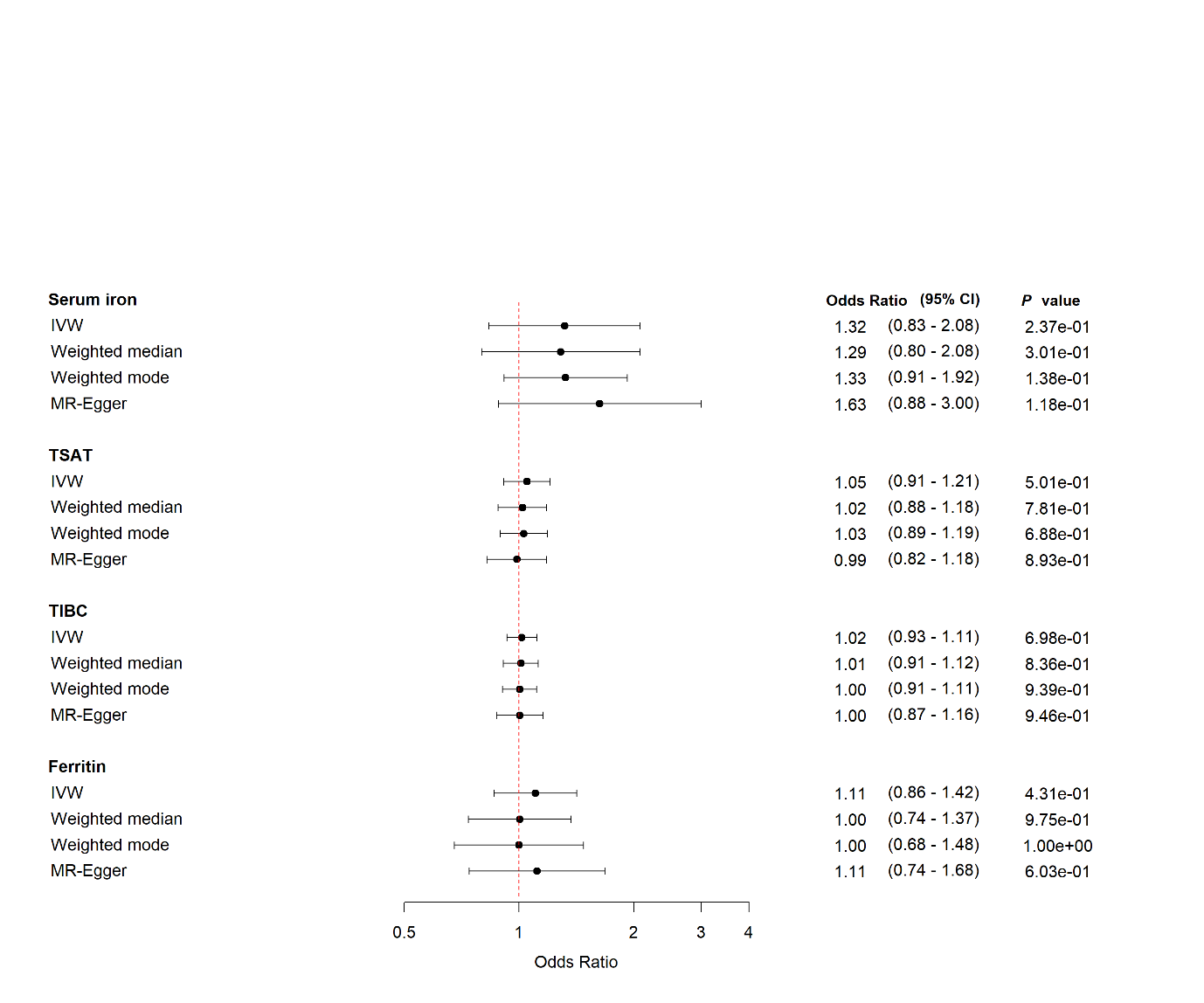

CI: Confidence Interval, Tsat; transferrin saturation, TIBC; total iron binding capacity

**Supplemental Figure S5: Forest plot for *women* with MR estimates for risk of being hospitalized with COVID-19 compared with non-hospitalized population**

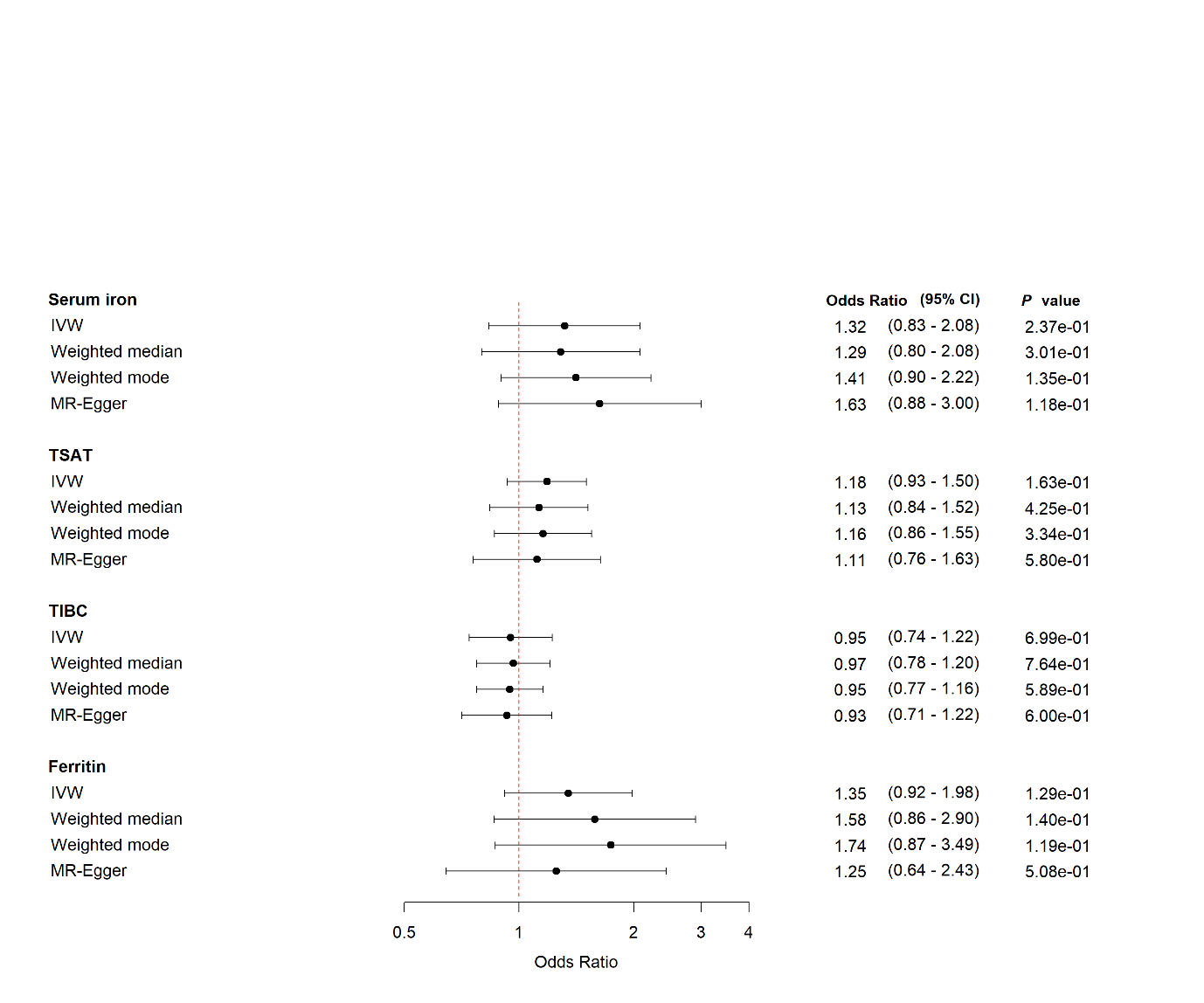

CI: Confidence Interval, Tsat; transferrin saturation, TIBC; total iron binding capacity

**Supplemental Figure S6: Forest plot for *men* with MR estimates for risk of being hospitalized with COVID-19 compared with non-hospitalized population**

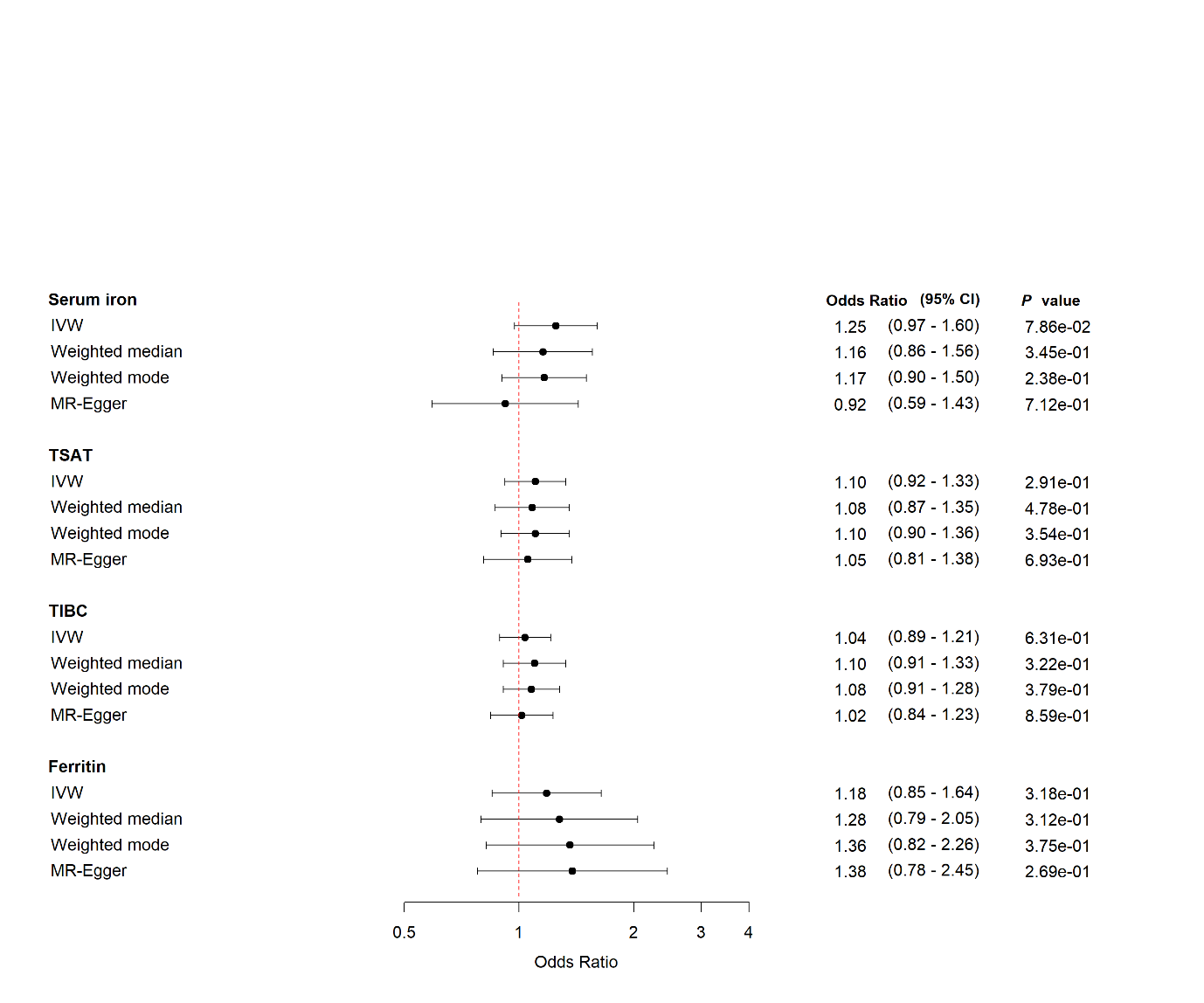

CI: Confidence Interval, Tsat; transferrin saturation, TIBC; total iron binding capacity
